# Phase I assessments of first-in-human administration of a novel malaria anti-sporozoite vaccine candidate, R21 in matrix-M adjuvant, in UK and Burkinabe volunteers

**DOI:** 10.1101/19009282

**Authors:** Navin Venkatraman, Alfred B. Tiono, Georgina Bowyer, Jonathan Powlson, Katharine A. Collins, Sam Coulibaly, Mehreen Datoo, Daniel Silman, Alphonse Ouedraogo, Issa Nébié, Egeruan Imoukhuede, Florian Brod, Pedro Folegatti, Emma Dickinson, Sophie Jamieson, Edith C. Bougouma, Daniel Wright, Duncan Bellamy, Amidou Diarra, Carly M. Bliss, Richard Morter, Greg Glenn, Louis F. Fries, Jenny Reimer, Karin Lovgren-Bengtsson, Megan Baker, Ian Poulton, Sarah Moyle, Eleanor Berrie, Nicola Green, Ekta Mukhopadhyay, Nicola Viebig, Brian Angus, Alison Lawrie, Rachel Roberts, Sarah C. Gilbert, David J.M. Lewis, Sodiomon B. Sirima, Katie J. Ewer, Adrian V. S. Hill

## Abstract

**Background:** Improvements in malaria control have stalled recently and new tools are needed. The R21 vaccine is comprised of the malaria circumsporozoite protein fused to hepatitis B surface antigen (HBsAg). It forms particles that lack the excess HBsAg in the frequently tested malaria vaccine candidate, RTS,S/AS01_B_.

**Methods:** We conducted an open-label, first-in-human, Phase Ia study evaluating safety and immunogenicity of R21 administered alone and with the saponin-based adjuvant, Matrix-M^™^ (MM). Twenty-eight healthy adults received three doses of R21 given intramuscularly 4 weeks apart. We subsequently conducted a Phase Ib randomised, controlled trial in West African adults.

**Findings:** Vaccinations were well tolerated, and the majority of local and systemic adverse events were mild. Reactogenicity was significantly lower in Burkinabe than UK vaccinees (p<0.0001). Antibody responses increased significantly 28 days after the 2^nd^ vaccination in UK volunteers. Antibody responses to R21 in all dose groups (2μg, 10μg and 50μg) were comparable to those of 50μg RTS,S/AS01_B_ in malaria-naïve adults at 28 days after final vaccination. The 10μg dose induced more durable responses, with 2-fold higher NANP-specific IgG titres at 6 months compared with the 2μg and 50μg dose groups. R21 also boosted baseline humoral responses in Burkinabe adults with well-maintained responses suggesting natural boosting.

**Interpretation:** R21 adjuvanted with MM is safe and has comparable immunogenicity to RTS,S/AS01_B_, even when administered at a five-fold lower 10μg dose in UK and African populations. This forms the basis for efficacy testing of this vaccine which could prove to be particularly cost-effective to manufacture and deploy.

## INTRODUCTION

Malaria remains one of the leading infectious causes of mortality worldwide and is caused by four species of the Plasmodium parasite in humans. *Plasmodium falciparum* predominantly affects children and pregnant women in sub-Saharan Africa and caused an estimated 219 million cases of malaria worldwide in 2017 (1). Increases in the distribution of long-lasting insecticidal nets (LLINs), widespread deployment of rapid diagnostic tests to target treatment with artemisinin-based combination therapy and use of intermittent preventive treatment in pregnant women have all contributed to a reduction in cases since 2010 (2). However, progress has been highly variable, with 15 countries in sub-Saharan Africa and India experiencing nearly 80% of malaria cases and since 2016, cases have increased in the 10 African countries with the greatest disease burden (1). Recent advances in control interventions include seasonal malaria chemoprophylaxis (SMC) programs with nearly 16 million children receiving treatment in 2017 (3). The emergence and spread of resistance to artemisinins (4, 5) and insecticides (6), threatens malaria control efforts even further and so there remains an urgent need for a durable, efficacious malaria vaccine.

The most advanced malaria vaccine candidate, RTS,S/AS01_B_, designed in 1987, elicits antibodies to the pre-erythrocytic circumsporozoite protein (CSP) and has completed testing in a large Phase III clinical trial (7-10). The World Health Organisation’s (WHO) Strategic Advisory Group of Experts (SAGE) on Immunization and the Malaria Policy Advisory Committee (MPAC), recommended further evaluation in a Malaria Vaccine Implementation Programme (MVIP), which has started in Malawi, Ghana and Kenya co-ordinated by WHO (11, 12). Though RTS,S has demonstrated significant vaccine efficacy of 46% in children after three doses (7), this falls short of the goals set down by the Malaria Vaccine Technology Roadmap - development of a suitable vaccine with at least 75% durable efficacy against clinical malaria by 2030 (13).

CSP, which is pivotal in sporozoite development and invasion of hepatocytes, has long been considered as one of the leading pre-erythrocytic antigens for the development of a malaria vaccine. R21 has been designed and developed at the Jenner Institute, University of Oxford since 2012 (14). This is an improved RTS,S construct, containing recombinant particles expressing the central repeat and the C-terminus of the CSP fused to the Hepatitis B surface antigen (HBsAg). It does not contain the four-fold excess of unfused HBsAg protein present in RTS,S, which was required for particle formation(15). CSP comprises around 20% of the total protein content in RTS,S and a large proportion of the antibody response induced by RTS,S is towards the HBsAg (16). In contrast, R21 contains only fusion protein moieties, with no unfused HBsAg, which increases the density of CSP antigen on the virus-like particle (VLP). As a result, a 50μg dose of R21 contains about 25μg of CSP antigen compared with 10μg of CSP in a standard adult 50μg dose of RTS,S. This was achieved by expressing R21 in an improved yeast expression system, Pichia pastoris, rather than in Saccharomyces cerevisiae. At the C-terminus of R21, a four amino acid sequence was added, EPEA (C-tag), for efficient immunochromatographic purification of R21. This sequence is found many times in the proteome of malaria parasites and humans but had not, to our knowledge, been used previously in any vaccine administered to humans (17).

In murine studies, R21 predominantly induced antibodies to CSP rather than HBsAg with comparable CSP immunogenicity and efficacy to RTS,S. Sporozoite challenge (1000 sporozoites per mouse injected intravenously) using transgenic P. berghei parasites (expressing the Plasmodium falciparum CSP homologue) performed in mice showed that R21 adjuvanted with Matrix-M™(MM) elicited 87.5 to 100% sterile protection (14).

Adjuvants can enhance the immunogenicity and efficacy of protein or VLP-based vaccines, and MM is an attractive relatively novel vaccine adjuvant for this purpose. As with other matrix formulations of Quillaja saponins, it shows acceptable safety in large numbers of recipients (18), and the ability to enhance both cellular and humoral immune responses to a range of antigens (19-24). In addition to the saponin, QS21, the AS01 adjuvant used with RTS,S also contains 50 μg of the immunostimulant, monophosphoryl lipid A (MPL). No such TLR4 ligand is present in the MM adjuvant which could lead to an improved safety profile and lower costs of manufacture. Therefore, we initially conducted a Phase I, first-in-human, open-label clinical trial to assess the safety and immunogenicity of R21 administered alone and with the novel saponin-based adjuvant, MM, in healthy UK volunteers. On the basis of an encouraging safety profile and comparable humoral immunogenicity to RTS,S/AS01_B_, but at the lower dose of 10μg, we tested this dose in Burkinabe adult volunteers and extended the UK study to assess the immunogenicity of an even lower dose of 2μg R21.

## METHODS

### Study design and participants

The Phase Ia study was conducted in healthy adults between the ages of 18 and 50 years at the Centre for Clinical Vaccinology and Tropical Medicine at the University of Oxford and the NIHR Imperial Clinical Research Facility, London in the United Kingdom.

Subsequently, the Phase Ib study was conducted in healthy Burkinabe adults aged between 18 and 45 years at the Centre National de Recherche et Formation sur le Paludisme (CNRFP) research unit, Banfora in Burkina Faso. Details on trial site in Burkina are provided elsewhere (25). All participants provided written informed consent. The Phase Ia study was an observational, open-label, first-in-human clinical trial of R21 at a range of doses. Eligible participants were assigned to one of four groups and received three doses of R21 at either 2μg, 10μg or 50μg administered alone or formulated with Matrix-M (Figure 1). The first three vaccinations in Groups 1-3 occurred in a staggered manner and interim safety reviews were conducted by the local safety monitor prior to progression to the higher dose group. The Phase Ib study was a single-blinded randomised, controlled trial assessing three doses of R21 10μg administered with MM in Burkinabe adults against a saline placebo (Figure 1). Despite single-blinding, investigators were unaware of group assignment throughout the study duration. The Phase 1b study only commenced after a satisfactory Data Safety and Monitoring Board (DSMB) review of the interim safety report for the 10μg and 50μg dose of R21 given to volunteers in the Phase Ia study. Volunteers were randomised to receive R21/MM or normal saline placebo. Full details regarding the study conduct are provided in the protocols for the two studies, which are available online (26, 27).

**Figure 1.**
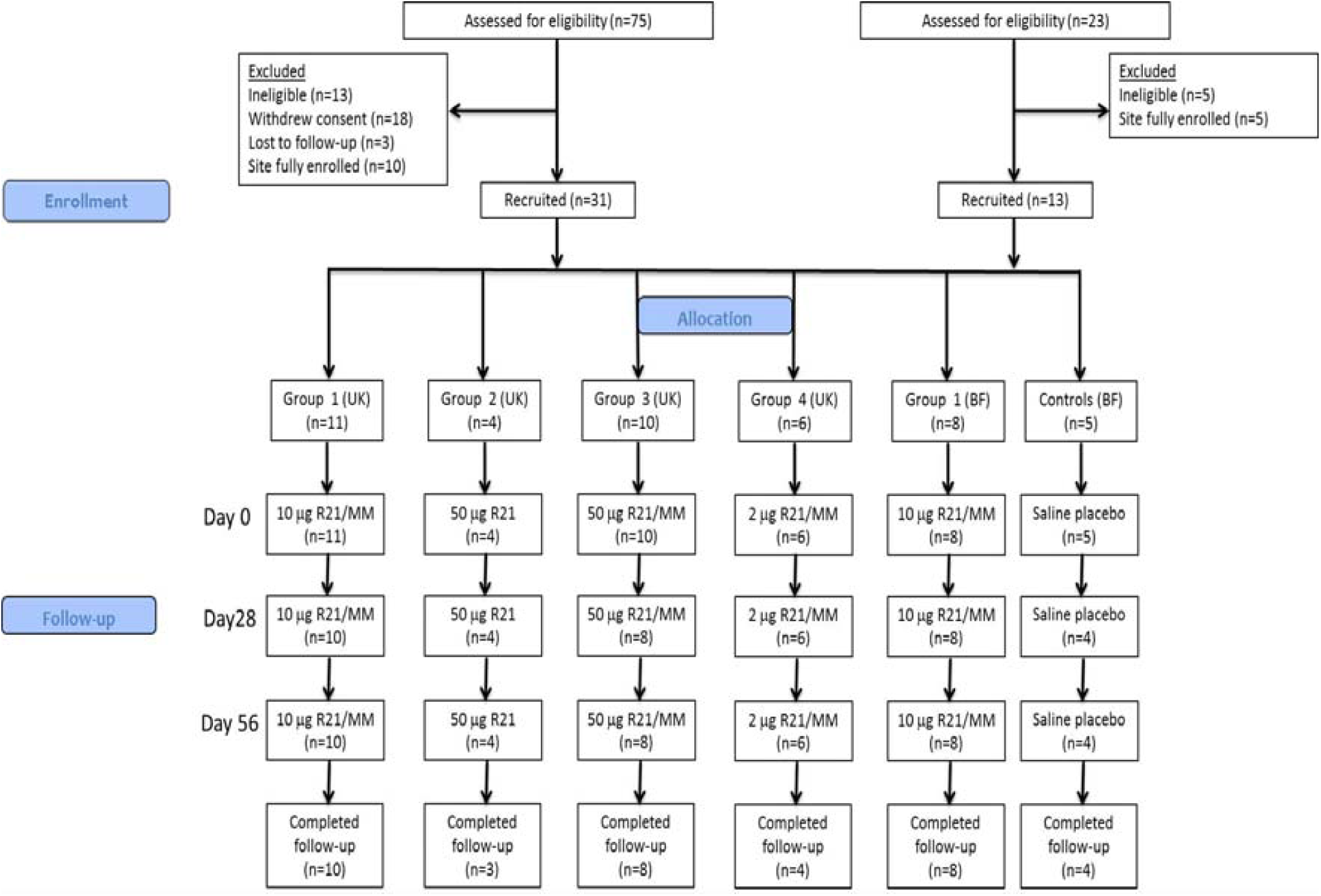
Flow chart of study design and volunteer recruitment. Abbreviations: BF, Burkina Faso; MM, Matrix-M; UK, United Kingdom. A total of 75 volunteers were screened and 31 were enrolled in total in the UK trial. One Group 1 volunteer withdrew consent after their first vaccination and was subsequently replaced. One Group 2 volunteer withdrew after their third vaccination and two Group 3 volunteers withdrew after their first vaccination and were not replaced. A total of 25 volunteers completed the UK study. A total of 23 volunteers were screened and 13 volunteers were enrolled in total in the BF trial. One volunteer in the placebo group was lost to follow up after the first dose and was replaced.

The Phase Ia study protocol and associated documents were reviewed and approved by the UK National Research Ethics Service, Committee South Central–Berkshire B (Ref: 15/SC/0386), the Medicines and Healthcare Products Regulatory Agency (Ref: 21584/0352/001-0001), and the Oxford University Clinical Trials and Research Governance team, who independently monitored compliance with Good Clinical Practice guidelines. The trial was registered with ClinicalTrials.gov (Ref: NCT02572388) and an independent local safety monitor provided safety oversight. The study protocol for the Phase Ib study was approved by the Burkina Faso regulatory authorities, The Burkina Faso Ministry of Health Ethical Committee for Biomedical Research (2014-10-118), the institutional review board of the Centre National de Recherche et de Formation sur le Paludisme (CIB/CNRFP), and Oxford Tropical Research Ethics Committee (OXTREC Reference: 36-15). The trial was registered with ClinicalTrials.gov (Ref: NCT02925403). An independent DSMB provided oversight and reviewed preliminary safety data before vaccinations commenced. The trial was monitored by an external organization (Margan Clinical Research Organization). Both studies were conducted according to the principles of the Declaration of Helsinki (2008) and the International Conference on Harmonization Good Clinical Practice guidelines.

### Procedures

R21 was thawed to room temperature and administered intramuscularly into the deltoid muscle within 1 hour of removal from the freezer, either alone, or mixed with an adjuvant at the bedside, immediately prior to administration.

Volunteers were observed for 60 minutes following vaccine administration. In the Phase Ia study, follow-up visits were scheduled for days 1, 7, 14, 29, 35, 42, 57, 63, 70, 84 and 238, with an additional visit at day 3 post-vaccination for the first three volunteers in Groups 1-3.

All participants recorded their temperature and any solicited local and systemic adverse events for 7 days post-vaccination and unsolicited adverse events for 28 days post-vaccination using an electronic diary. A review of solicited and unsolicited adverse events (AEs) occurred at each follow-up visit. Safety bloods including full blood count, renal function and liver function tests were done on visits at day 0, 7, 28, 35, 56, 63, 84 and 238. In the Phase Ib study, volunteers were visited at home daily for 6 days after each vaccination by a field worker for assessment and recording of any solicited and unsolicited AEs in diary cards. They were also seen in clinic at day 7 and day 28 after each vaccination and attended a final follow up visit one year after enrolment. Safety bloods including full blood count, creatinine and alanine aminotransaminase were done in clinic at day 0, 7, 28, 35, 56, 63, 84, 140 and 365. Severity grading of AEs and the assignment of a causal relationship for AEs were conducted according to predefined guidelines stated in the protocol, which were harmonised across both clinical trials for grading of solicited AEs. For unsolicited AEs, MedDRA (MedDRA® the Medical Dictionary for Regulatory Activities terminology) was used for coding in the UK clinical trial. In the Burkinabe trial, the DAIDS AE grading table was used (28).

Antibody responses measured by anti-NANP IgG ELISA were performed at all time points and IgG antibody avidity was assessed by sodium thiocyanate (NaSCN)-displacement ELISA. To assess whether antibodies to the C-tag used for R21 purification were induced by vaccination, N-terminal biotinylated peptides were constructed for the C-tag (EPEA), the C-tag plus the four adjacent amino acids in the R21 construct (WVYIEPEA) forming an 8-mer and the C-tag plus the eleven adjacent amino acids in the R21 construct (LPIFFCLWVYIEPEA) forming a 15-mer. For hepatitis B, antibodies to the HBsAg were measured using the Abbot Architect 2000i chemiluminescent micro-particle immunoassay (CMIA). Antibody concentration of 100.0mIU/ml or greater was considered positive. Only groups 1, 2 and 3 were assayed for C-tag antibodies. Ex-vivo IFN-γ ELISpot responses to CSP were assessed on samples from day 0, 42 and 84, (ELISpots were not performed in the Burkinabe cohort as PBMC were not collected). Full details of the methods for all assays are provided in the supplementary appendix.

### Outcomes

The primary objective was to assess the safety and tolerability of R21 with and without adjuvant MM in healthy UK and Burkinabe volunteers. The primary outcome measures included occurrence of solicited local and systemic reactogenicity signs and symptoms for 7 days following the vaccination, occurrence of unsolicited AEs for 28 days following the vaccination, change from baseline for safety laboratory measures and occurrence of serious AEs during the whole study duration. The secondary objective was to assess the cellular and humoral immunogenicity of R21 with and without adjuvant MM in healthy UK volunteers and humoral immunogenicity in Burkinabe volunteers. The IgG response to the NANP repeat region was the primary immunogenicity readout, as this measure is associated with efficacy of RTS,S/AS01_B_ (29).

### Statistical analysis

Data were analysed using GraphPad Prism version 8.11 for Mac (GraphPad Software Inc., California, USA) and Stata 10.0 (Statacorp LP, Texas, USA). Medians with interquartile ranges for each group are described. Kruskal-Wallis analysis and the Friedman test were used to compare peak immune responses with baseline. Significance testing of differences between two groups used Mann-Whitney analysis. A Wilcoxon matched-pairs analysis was used to compare between time points within groups. A chi-squared test for trend was used to compare the safety data between different groups. A value of p < 0.05 was considered statistically significant.

## RESULTS

### Study Populations

The Phase 1a study in the UK was performed between the 1^st^ October 2015 and 3^rd^ January 2017, and 31 of the 75 volunteers who were screened for eligibility were enrolled in the UK study. One volunteer in Group 1 withdrew after their first vaccination and was replaced. One Group 2 volunteer withdrew after their third vaccination. Two volunteers in Group 3 withdrew after their first vaccination and were not replaced, at withdrawal there were no ongoing AEs and safety bloods were normal. All remaining volunteers completed follow-up. The Phase 1b study in Burkina Faso was performed from 26^th^ August 2016 to 28^th^ September 2017. 13 of the 23 participants who were screened for eligibility were enrolled. Eight volunteers completed follow-up after receiving three doses of 10μg R21/MM in addition to four volunteers who received a saline placebo. One participant in the placebo group was lost to follow-up after the first dose and was replaced. Participant flow in the two clinical trials is summarised in Figure 1 and demographic data are summarised in Table 1.

**Table 1.** Baseline characteristics of enrolled volunteers. Mean (SD) age for each cohort is shown

|  | UK cohort (n=31) | Burkina Faso cohort (n=13) |
| --- | --- | --- |
| **Sex** |  |  |
| Male | 15 (48%) | 10 (77%) |
| Female | 16 (52%) | 3 (23%) |
| **Age (years)** | 29.9 (8.3) | 30.5 (8.1) |

### Adverse events

No serious adverse reactions (SARs) or suspected unexpected serious adverse reactions (SUSARs) occurred. Two SAEs occurred; the first was deemed unlikely and the second not related to vaccination. Solicited local and systemic AEs in the first 7 days after each vaccination related to R21 vaccination in the UK population are shown in Figure 2 and in Supplementary Table 1. The majority of AEs reported were mild in severity and self-limiting. As expected, the addition of MM increased the reactogenicity of the 50μg dose compared with administration of 50μg of R21 alone. There was a significant trend for more reactogenicity in the higher dose groups than in the very low dose 2μg group (p<0.0001, Chi-squared test for trend across doses) where minimal reactogenicity was observed. Vaccine site pain was the most common local AE and was predominantly mild in severity. One volunteer in Group 1 reported a mild fever (37.7°C) and another volunteer in Group 1 reported a moderate fever (38.1°C). Both occurred after the 2^nd^ vaccination and resolved within 24 hours. One volunteer in Group 2 reported a fever of 39°C associated with a constellation of flu-like symptoms, eight days after their first vaccination, which resolved within 24 hours. Severe AEs were only reported in the 50μg R21/MM group by three volunteers and resolved within 48 hours.

**Figure 2.**
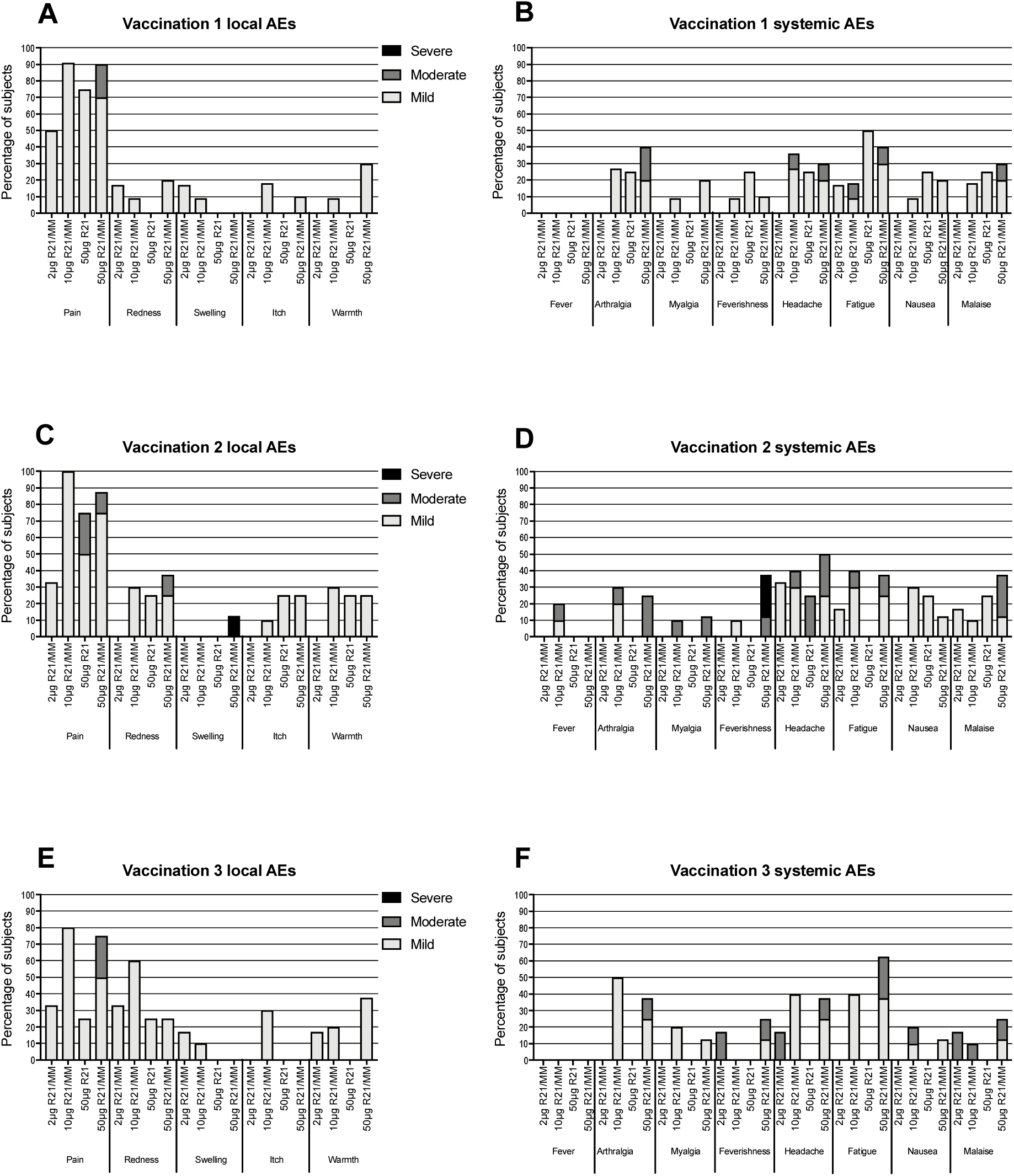
Local (A, C, E) and systemic (B, D, F) solicited adverse events (AEs) reported by UK volunteers in the electronic diary card during the first seven days related to each vaccination given 4 weeks apart. R21 was either administered alone (n=4) or mixed with Matrix-M at different doses of 2µg (n=6), 10µg (n=10) and 50µg (n=8). Only the highest intensity of each AE per subject is listed. Data are combined for all AEs for all volunteers receiving the same vaccine at the stated time point.

Local and systemic AEs to 10μg R21 adjuvanted with MM in Burkinabe adults are shown in Figure 3. Very few solicited systemic AEs were reported and most local AEs were mild in nature with overall reactogenicity was significantly reduced compared to UK volunteers receiving the same dose, summarised in Table 2 and S2 (p<0.0001, Chi-squared test). No volunteers reported severe AEs. Vaccine site pain was again the most commonly reported local AE. There were no episodes of fever associated with vaccination in the Burkinabe cohort. No solicited AEs were recorded in the saline placebo group.

**Table 2.** Summary of adverse event (AE) frequency in UK and Burkina Faso (BF) cohorts immunised with 10μg R21/MM and between two UK cohorts immunised with either 50μg R21/MM or 50μg RTS,S/AS01B (data from (30)). The total number of local and systemic AEs reported by all volunteers following all vaccinations in each vaccine regime is shown (Pearson’s chi-squared test for trend). *, comparison with group immunized with 50µg R21/MM.

|  | All AEs all vaccinations |  |  |  |  |
| --- | --- | --- | --- | --- | --- |
| Location | UK | BF | UK | UK | UK |
| Vaccine/Adjuvant | R21/MM | R21/MM | RTS,S/AS01<sub>B</sub> | R21/MM | R21 alone |
| Dose Vaccine | 10µg | 10µg | 50µg | 50µg | 50µg |
| N enrolled | 11 | 8 | 17 | 10 | 4 |
| No. AEs |  |  |  |  |  |
| Mild | 94 | 5 | 166 | 65 | 19 |
| Moderate | 9 | 5 | 83 | 26 | 2 |
| Severe | 0 | 0 | 23 | 3 | 0 |
| Total AEs | 103 | 10 | 272 | 94 | 21 |
| Chi-Squared (Pearson) | p<0.0001 |  | p<0.0001 |  | P=0.002* |

**Figure 3.**
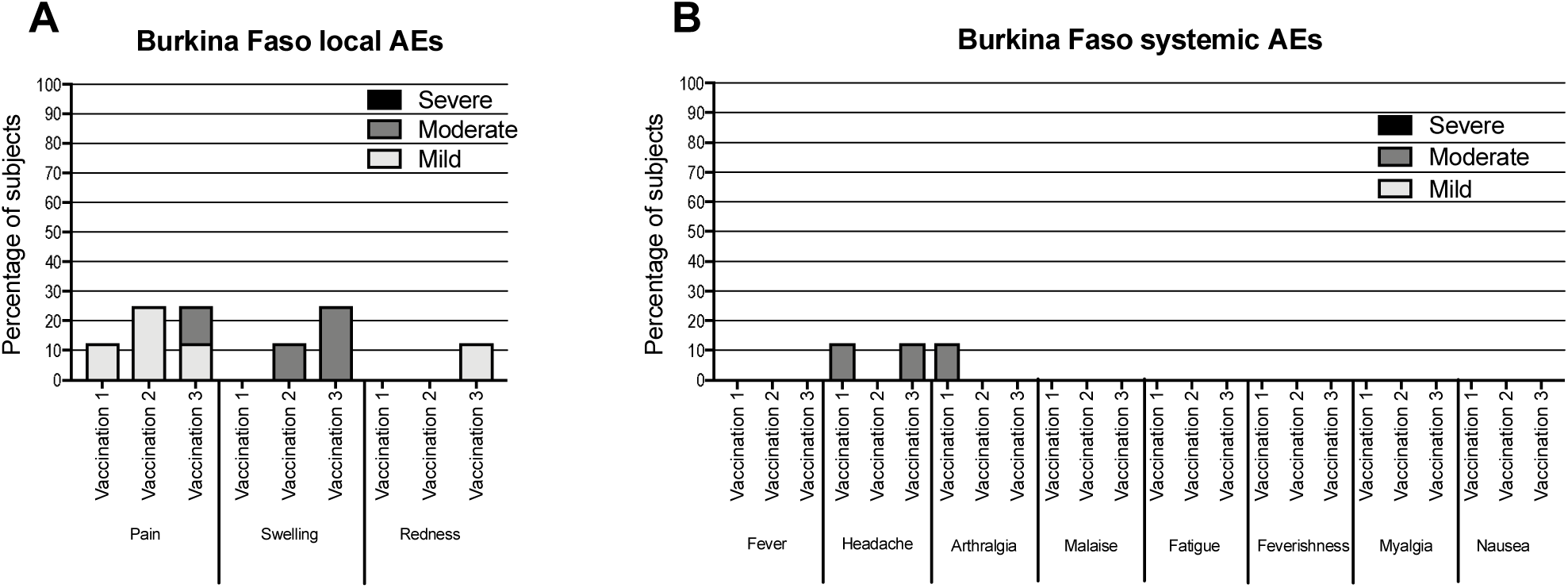
Local and systemic solicited adverse events (AE) reported by Burkinabe volunteers (n=8) during the first seven days related to vaccination with a dose of 10µg R21/MM. Only the highest intensity of each AE per subject is shown. Data are combined for all AEs for all volunteers receiving the same vaccine at the stated time point. No solicited AEs were observed in the saline placebo group.

Furthermore, the reactogenicity profile observed in the 50μg R21/MM group is significantly reduced (p < 0.0001, Pearson’s chi-squared test for trend) compared to that observed in a previous clinical trial conducted in our institute using the same AE grading criteria, where volunteers received three doses of 50μg RTS,S/AS01_B_ given four weeks apart (30). This was mainly due to a significant reduction in the number of systemic AEs reported with a higher incidence of moderate and severe AEs reported by volunteers receiving RTS,S/AS01_B_ (Table 2). In particular there were no post-vaccination fevers in the 50μg R21/MM group (0% compared with 26% for 50μg RTS,S, p=0.004 chi-squared test).

Unsolicited AEs collected for 28 days after each vaccination and those deemed possibly, probably and definitely causally related to vaccination were predominantly mild in nature for the UK trial (Table S3). Laboratory AEs were predominantly Grade 1 in the UK cohort and are tabulated in Table S4. In the Burkinabe cohort, all unsolicited AEs were deemed unlikely or unrelated to vaccination (Table S5). A higher frequency of laboratory AEs were observed in the Burkinabe cohort in both the R21/MM and saline placebo vaccinated groups (Table S6).

### Immunogenicity

#### Humoral responses

IgG antibody titres to the NANP repeat region of the CSP antigen were measured by ELISA in the same laboratory by the same operator for both trials. In the UK cohort, addition of the Matrix-M adjuvant markedly increased antibody levels compared with R21 administered alone. Increasing titres of NANP-specific IgG were detected with increasing doses of R21 after the first and second vaccinations, however there was no significant difference between dose groups after the third immunisation (Kruskal-Wallis analyses, p=0.02, p=0.04 and p=0.12 at D28, D56 and D84, respectively, Figure 4A and C). At day 238, titres were higher in the 10μg dose group than in the 50μg and 2μg groups (Kruskal-Wallis test with Dunn’s multiple comparison test, p=0.06, Figure 4A and C).

**Figure 4.**
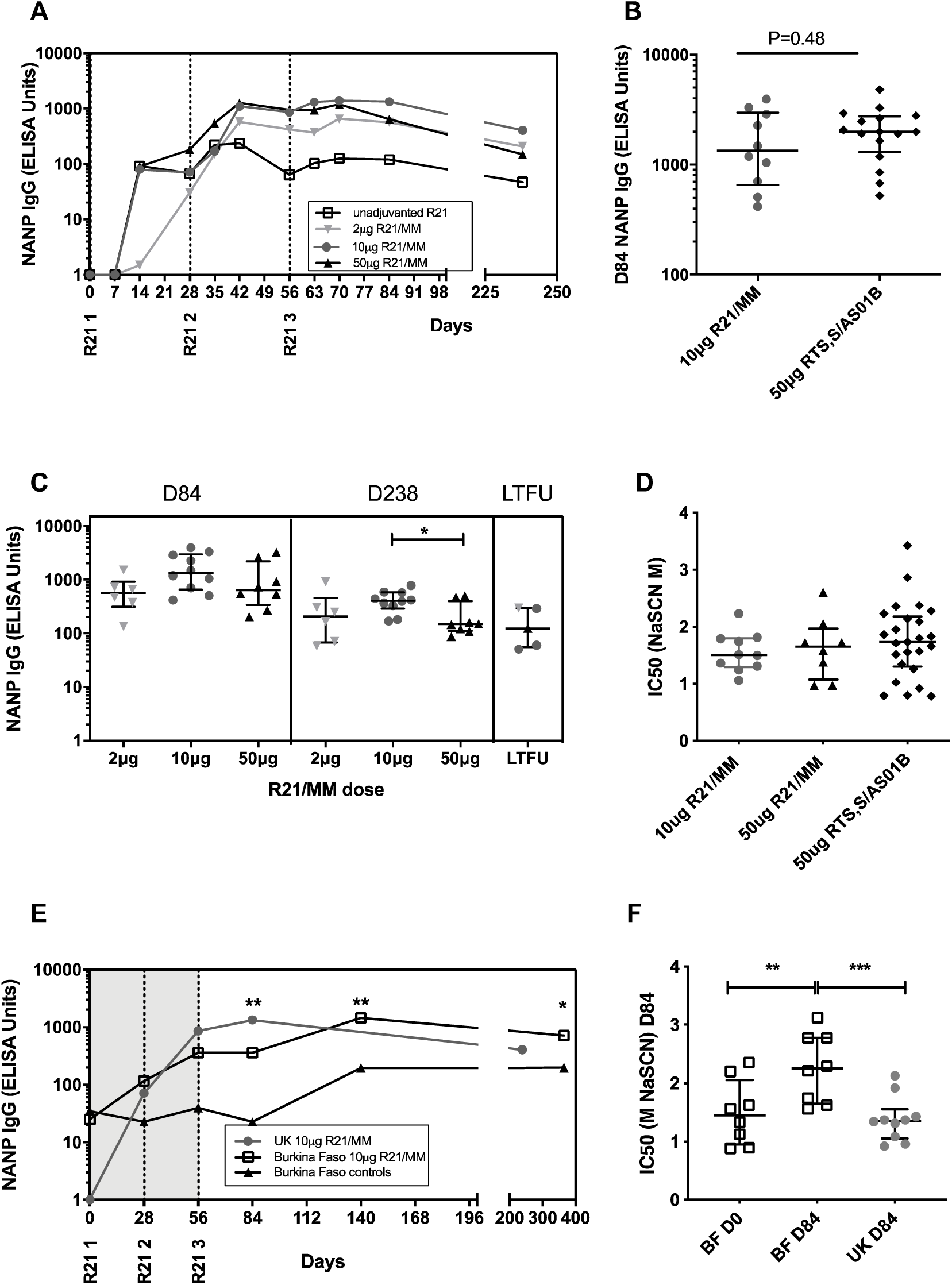
Antibody responses to R21/MM. A) Median NANP IgG time courses in UK adults vaccinated with 10µg R21/MM (G1, circles, n=10), 50µg unadjuvanted R21 (G2, squares, n=3) or 50µg R21/MM (G3, triangles, n=8). B) Peak NANP IgG titres (D77-D84) in 10µg R21/MM group and in groups from two previous trials that were vaccinated with three doses of 50µg RTS,S/AS01B, Mann-Whitney p=0.48 (1, 2). C) Individual NANP IgG responses at peak (D84) and D238 in UK adults by dose and a long-term follow-up visit (LTFU) between 22 and 35 months after first vaccination. Mann-Whitney analyses between groups at D84 p=0.20 and D238 (p=0.02). Bars represent medians + interquartile ranges, IQRs. D) Avidity of NANP-specific IgG at peak (D84) in 10 and 50µg R21/MM groups and 50µg RTS,S/AS01B from two previous trials as in Figure 4B, Kruskal-Wallis p=0.08. E) NANP IgG time courses (median + IQRs) in UK adults in G1 (10µg R21/MM, circles) and Burkina Faso adults given the same dose of R21/MM (squares) or placebo (triangles). Titres at each time point were compared by Mann-Whitney analysis: D28 p=0.75, D56 p=0.12, D84 p=0.02 (UK vs. Burkinabe, R21/MM). Between D84 and D140, tires increased significantly in the Burkabe cohort (Wilcoxon matched-pairs 2-tailed, p=0.008) and at both D140 and D365, titres in the R21/MM group remained significantly higher than in the placebo control group (Mann-Whitney test, p=0.004 D140, p=0.04 D365). F) Antibody avidity at baseline and peak immune response (D84) in 10µg R21/MM groups (Mann-Whitney, p=0.008 for BF D0 comparison with D84, p=0.001 for D84 comparison between UK and BF).

Based on the quantity of CSP contained in each vaccine, the 10μg dose of R21 is most similar to the standard 50μg dose of RTS,S. IgG titres were compared to those in UK participants receiving three 50μg doses of RTS,S in previous UK clinical trials at the Jenner Institute.

ELISA assays were run using the same protocol in the same laboratory by the same operator. There was no evidence of a difference between the peak IgG response to three 10μg doses of R21 in MM to those induced by three 50μg doses of RTS,S/ AS01_B_ (Mann-Whitney test, p=0.5, Figure 4B) (30, 31). Avidity of IgG antibodies at the peak of the immune response did not vary between either dose of R21 in MM or compared to vaccination with RTS,S (Kruskal-Wallis test, p=0.6, Figure 4D).

In the Burkinabe cohort, pre-vaccination titres to CSP were higher than in UK participants due to malaria exposure. Responses to vaccination did not differ significantly between the two cohorts after the first two vaccinations, however the third dose of R21 failed to boost responses in the Burkinabe cohort, and as a consequence, titres were significantly lower at D84, four weeks after the third dose (Mann-Whitney test, p=0.02, Figure 4E) compared to UK vaccinees. By D140, responses in the Burkinabe R21/MM cohort had increased significantly from D84 (Wilcoxon matched-pairs 2-tailed, p=0.008), and were significantly higher than those in the placebo group at D140 and D365 (Mann-Whitney test, p=0.004 D140, p=0.04 D365). Although titres were lower at D84 in the Burkinabe cohort than the UK, antibody avidity was significantly higher (Mann-Whitney test, p=0.001, Figure 4F). The avidity of the antibody response also increased significantly after vaccination in the Burkinabe cohort (Mann-Whitney test, p=0.008). The induction by R21/MM of high avidity antibodies, and initial evidence of boosting through natural malaria exposure in Burkinabe adults, with antibody titres comparable to those observed in UK vaccinees at D365, is interesting, particularly as it has been difficult to detect evidence of significant immune boosting by natural malaria exposure in RTS,S/AS01_B_ vaccinees in malaria-exposed or semi-immune adults (32, 33).

The functional activity of CSP-specific antibodies was assessed in vitro by measuring blocking of sporozoite invasion into a human hepatoma cell line in the presence of serum from vaccinees (inhibition of sporozoite invasion, ISI assay). Prior to vaccination, serum from UK participants did not demonstrate ISI activity, while serum from Burkinabe volunteers demonstrated up to 50% inhibition (p=0.02, Mann-Whitney, Figure 5A). Vaccination with R21/MM significantly increased the ISI activity from baseline to D84 in the UK cohort (p=0.03, Wilcoxon matched-pairs) and there was a trend towards higher ISI activity in the Burkinabe participants at D84 than in the UK participants (p=0.06, Mann-Whitney). An association was detected between antibody titre and ISI activity in the Burkinabe cohort (r=0.81, p=0.07, Spearman’s rank test, Figure 5B), but not in the UK cohort.

**Figure 5.**
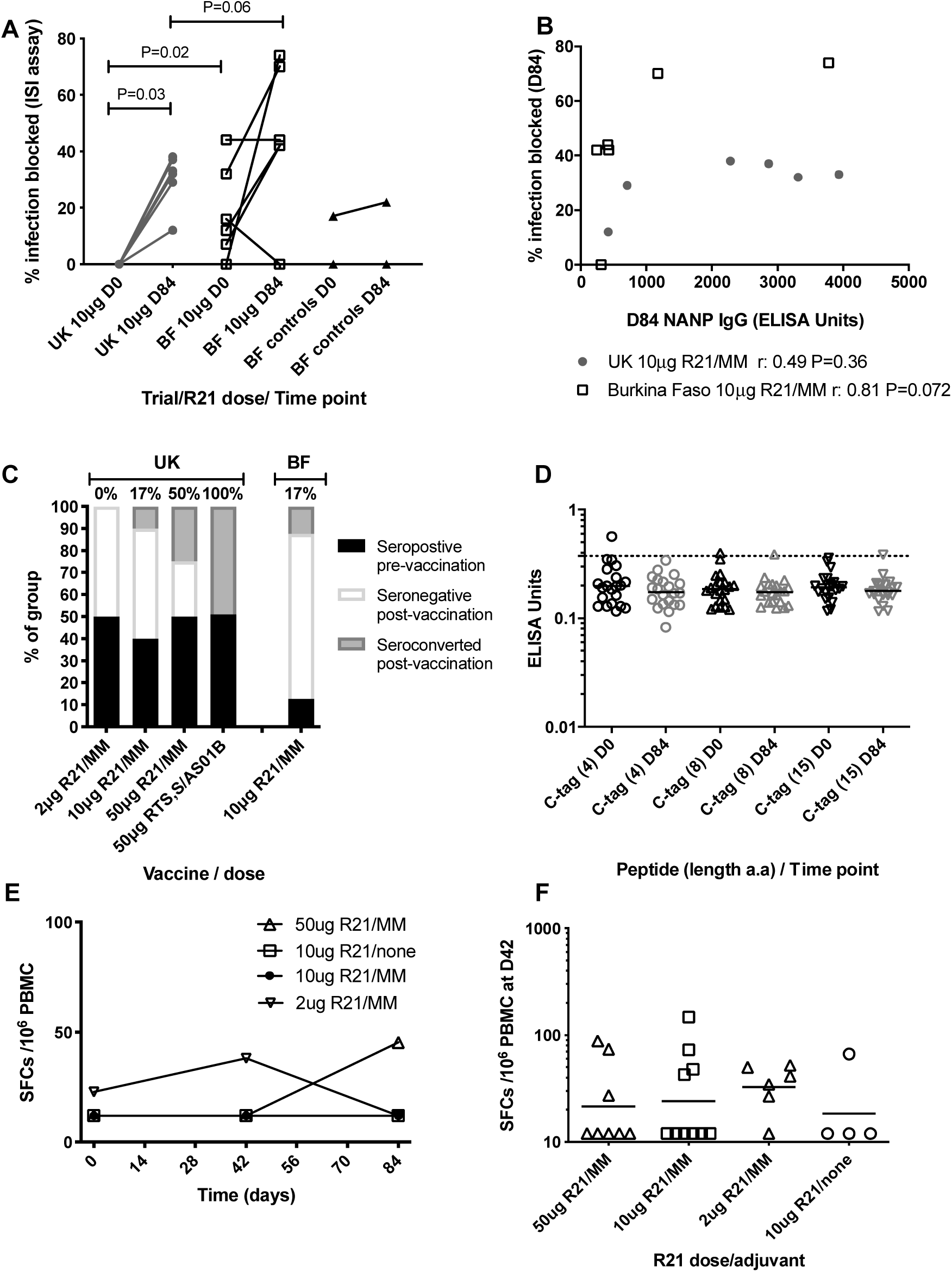
Extended R21/MM immunology. A) Percentage of infection blocked in the inhibition of sporozoite invasion assay (ISI) at baseline and D84 in a subset of UK and Burkinabe adults vaccinated with 10µg R21/MM and Burkinabe adults vaccinated with saline (controls). Wilcoxon matched-pairs analyses for change over baseline and 2-tailed Mann-Whitney for comparisons between groups). B) Association between D84 NANP IgG titres and the percentage of infection blocked in UK and Burkinabe adults vaccinated with 10µg R21/MM. Spearman rank analyses. C) Hepatitis B surface antigen antibody responses in each group before and after vaccination. D) ELISA responses to C-tag peptides at day 0 (pre-vaccination) and Day 84 (4 weeks post-vaccination). Line shows cut-off for positivity, determined as the mean responses at day 0 plus three standard deviations. E) Median CSP-specific T cell responses as measured by IFNγ ELISpot. F) individual CSP-specific T cell responses for each group at the peak time point (day 42).

Between 40% and 50% of participants in each UK group were seropositive to HBsAg prior to R21 vaccination, presumably due to previous vaccination against Hepatitis B. Of those that were seronegative at vaccination, 0, 17% and 50% of volunteers seroconverted for HBsAg after 3 doses of R21/MM in the 2μg, 10μg and 50μg dose groups, respectively. This is in contrast to vaccination with 3 doses of RTS,S/AS01_B_ which induced seroconversion in 100% of seronegative vaccinees in a Belgian study (34) (Figure 5C). In the Burkinabe cohort, seroconversion was detected in only 1 of 7 (17%) volunteers that were seronegative prior to vaccination.

A four amino acid label (EPEA) was added to the C-terminal of the R21 construct (C-tag) to facilitate protein purification during biomanufacture. Antibody response to the C–tag was assessed by ELISA in the UK cohort only. C-tag IgG responses were not induced by vaccination with R21, although one participant had a very weak response to EPEA pre-vaccination, which was not detected post-vaccination at Day 84 (Figure 5D).

#### Cell-mediated immunity

T cell responses to CSP were enumerated by ex vivo IFN-γ ELISpot and were weak in R21/MM vaccinees (Figure 5E) and undetectable in the unadjuvanted group. The magnitude of responses were comparable to those observed in recipients of RTS,S/AS01_B_ (35). Responses peaked at day 42, two weeks after the second dose and there were no significant differences between groups (Figure 5F).

## DISCUSSION

We report here the safety and immunogenicity of the first-in-human administration of the novel malaria vaccine candidate, R21, administered with the saponin-based adjuvant, MM in UK and African adults. This vaccination approach was safe and very well tolerated with very low doses inducing high antibody levels, comparable to those previously associated with protection in humans induced by other malaria anti-sporozoite vaccines. The reactogenicity profile of 50μg R21/MM was significantly improved compared to the standard adult 50μg RTS,S/AS01_B_ dose in healthy adult UK volunteers (30) and importantly, post-vaccination fever was not detected. An effective vaccine will be an essential tool in the malaria elimination and eradication efforts and the absence of fever is a significant benefit in the context of mass administration campaigns. Minimal reactogenicity was detected in African adults and this was significantly reduced compared to UK adults at a dose of 10μg R21/MM.

Humoral responses observed to the conserved central NANP repeat region are comparable to previous RTS,S/AS01_B_ data from two clinical trials conducted in the UK where the same ELISA was used. The humoral response is predominantly responsible for the efficacy of RTS,S/AS01_B_ against malaria infection (36). It is therefore encouraging that comparable humoral responses are elicited at 28 days after the third vaccination even at the very low dose of 2μg and 10μg. This could have significant dose-sparing and cost-saving implications for vaccine production.

R21/MM administered at doses of 10μg and 50μg did not result in significant boosting of the humoral response after the third vaccination compared to the peak response after the second vaccination. Though RTS,S is currently generally given in a three dose schedule, administered 4 weeks apart, preliminary studies with RTS,S assessed regimens where the third dose was delayed to 24-28 weeks (15, 37). Though these studies did not show significant boosting of the humoral response with a delayed third dose, higher level efficacy of 86% was observed in malaria-naïve adults who received RTS,S/AS02 given at 0, 1 and 7 months (37). However, these volunteers had also received one-fifth of the vaccine and adjuvant at the third administration due to earlier reactogenicity concerns. Studies in African adults living in malaria endemic regions have shown that there is boosting of the immune response after the third dose with a 0, 1 and 6 month schedule (38, 39). More recently, Regules et al. revisited a delayed fractional (i.e. one fifth) dose with RTS,S/AS01_B_ where high level efficacy was again observed. This was associated with increased somatic hypermutation and antibody avidity, rather than the magnitude of the humoral response (40).

Durable antibody responses were observed at 6 months after the final vaccination. The 10μg dose elicited significantly higher titres compared with the 50μg dose at 6 months. Though RTS,S/AS01_B_ demonstrated short-term efficacy in malaria-naïve and endemic populations, durable protective immunity is a hitherto unmet goal. It is well recognised that the antibody responses to the CSP elicited by RTS,S/AS01_B_ decay over time, which is reflected in reduced vaccine efficacy (33, 41, 42). That a lower dose of protein should induce a more durable response is of particular interest from an immunological perspective. In other settings, higher priming antigen doses favour production of antigen-specific plasma cells that only have a short lifespan, whereas lower doses can preferentially drive the induction of immune memory (43, 44). A few of these plasma cells differentiate into long-lived plasma cells (LLPC) and in the absence of subsequent antigen exposure; the proportion of LLPC generated by a vaccine is predictive of the durability of the antibody response (45). Antibody titres at 6-12 months after immunisation would reflect this as the initial short-term plasma cell response would decline.

A successful malaria vaccine is needed to help reduce malaria morbidity and mortality in over 30 million children born in malaria endemic regions of Africa each year, as well as for disease control and eventual eradication efforts in other continents. For a four-dose regimen, such as that under evaluation by MVIP, this would require at least 130 million doses per annum for infants in Africa alone. The finding that R21 can induce comparable immune responses to RTS,S using just a fifth to a 25^th^ of the dose used by the RTS,S suggests that substantial dose sparing should be possible reducing cost of goods and easing the challenge of making hundreds of millions of doses of vaccine each year. The initial improved safety profile of R21 observed with the Matrix-M adjuvant compared to the standard RTS,S/AS01_B_ regimen, along with good durability of immune response after the low dose R21/MM is also encouraging. Furthermore, R21 is now being manufactured by the Serum Institute of India, who are the world’s largest vaccine supplier by volume and supply 40% of the vaccines funded by GAVI (46), increasing the potential to manufacture R21/MM on the scale likely to be required for effective malaria control and eventual elimination.

In conclusion, these Phase I clinical trials showed that the novel malaria vaccine candidate, R21 adjuvanted with Matrix-M was safe and well tolerated in both UK and African subjects. Phase IIa studies to assess efficacy in an established malaria sporozoite challenge model are currently ongoing.

## Data Availability

All data related to this study is contained in the manuscript.

## ACKNOWLEDGEMENTS

We thank the members of the Data Safety Monitoring Board and all the study volunteers. The UK study was funded by the National Institute for Health Research (NIHR) Oxford Biomedical Research Centre (BRC). The views expressed are those of the authors and not necessarily those of the NHS, the NIHR or the Department of Health. Vaccine manufacture was funded by the UK Medical Research Council (Grant no: MR/JOO8680/1) and the European Commission FP7 programme (Grant no: 305282). The study in Burkina Faso was supported by a Strategic Primer grant award from the European and Developing Countries Clinical Trials Partnership (EDCTP) with co-funding from Swedish International Development Cooperation Agency (Sida); UK Medical Research Council; Irish Aid, Department of Foreign Affairs and Trade, Ireland; and Bundesministerium für Bildung und Forschung (BMBF), Germany. It was performed as part of the work programme of the EDCTP Malaria Vectored Vaccines Consortium 2 (MVVC2), (grant number SP.2011.41304.025). The European Vaccine Initiative (EVI) coordinated the EDCTP-funded MVVC project. Odile Leroy was executive director of EVI at the time of the study. N.K.V. is an employee of EVI. The work was also supported by the UK National Institute of Health Research through the Oxford Biomedical Research Centre (http://oxfordbrc.nihr.ac.uk/) (A91301 Adult Vaccine) and the Wellcome Trust (https://www.wellcome.ac.uk/) (084113/Z/07/Z). We are grateful to Shahid Khan and Chris Janse at LUMC for provision of transgenic parasite lines and to Alexandra Spencer, Ahmed Salman and Marta Ulaszewska for immunopotency assays and preparation of infected mosquitoes. We thank Philip Angell-Manning for his contribution to R21 manufacturing. We are also grateful to Ruth Payne and Saranya Sridhar for contributing to clinical care of participants and to Catherine Mair and Kate Harrison for assistance with sample processing. We thank Nicola Williams of the Nuffield Department of Primary Health Care Sciences at the University of Oxford for statistical review of the manuscript.

